# Investigation of Intra-Fraction Stability and Inter-Fraction Reproducibility of Deep Inspiration Breath-Hold Across Two Hypofractionated Radiotherapy Regimens in the HYPORT Adjuvant Study

**DOI:** 10.64898/2026.06.06.26355038

**Authors:** Anurupa Mahata, Debapriya Roy, Ramkrishna Khatua, Sougata Maity, Kasturi Barik, Santam Chakraborty, Jyotirmoy Chatterjee, Sanjoy Chatterjee

## Abstract

**Background:** Deep Inspiration Breath Hold (DIBH) is a widely used respiratory motion management technique for minimizing cardiac dose in left-sided breast radiotherapy. In the Breast HYPORT Adjuvant study, DIBH was employed for cardiac sparing in patients without nodal irradiation using a standardized institutional protocol with the Varian Real-time Position Management (RPM) system. Both moderate-hypofractionation (control arm - 40Gy in 15 fractions) and one-week hypofractionation (experimental arm - 26 Gy in 5 fractions) regimens were delivered using this protocol. This study aimed to evaluate the robustness of DIBH by analyzing intra-fraction stability and inter-fraction reproducibility of breath-hold amplitude across the two treatment regimens.

**Methods:** Respiratory waveforms acquired during each treatment session were analyzed to determine the median breath-hold amplitude and its standard deviation during beam delivery. Intra-fraction stability was assessed from variations within individual treatment sessions, while inter-fraction reproducibility was evaluated relative to the simulation waveform amplitude across all treatment sessions. These parameters were compared between the two HYPORT regimens to examine breath-hold consistency during treatment delivery. Moreover, an additional comparison was made between the one-week hypofractionation regimen and the first five fractions of the moderate-hypofractionation regimen to evaluate the effect of treatment duration . Lung volumes from free-breathing and DIBH CT scans were analyzed to assess the effectiveness of patient breath-hold training.

**Results:** Both arms demonstrated an average 1.7-fold increase of air volume in lung during the breath-hold position, confirming the effective implementation of DIBH during treatment planning and delivery. Structured training resulted in increased breath-hold amplitudes, with gains of 22.87% and 24.16% with respect to the first trial session in the experimental and control arms, respectively. Both regimens receive equivalent doses for approximately the same air volume in lung . Despite the different prescription doses in the two arms (26 Gy vs. 40 Gy), the experimental arm achieved an equivalent mean heart dose of 2.91% (75.6 cGy) compared with 2.95% (118.51 cGy) in the control arm, suggesting a similar cardiac preservation protocol adopted during treatment planning. Intra-fraction stability was similar between the control arm and the experimental arm, with median amplitude variations of 1.006 mm (95% CI: [0.998–1.015]) and 1.079 mm (95% CI: [1.067–1.097]), respectively. In contrast, inter-fraction reproducibility improved in the experimental arm, with lower deviation from simulation amplitude (0.44 ± 0.24 mm vs. 0.66 ± 0.25 mm) for the entire treatment schedule. The stability and reproducibility of experimental arm were further compared with the first five fractions of the control arm. The results were similar to those of the experimental arm.

**Conclusion:** In this study, we compared two treatment regimens in terms of intra-fraction stability and inter-fraction reproducibility during DIBH radiotherapy. Both regimens demonstrated comparable intra-fraction stability, indicating effective motion management irrespective of treatment duration. However, the experimental arm showed better interfraction reproducibility, suggesting more consistent breath-hold performance throughout the treatment course. Based on stability and reproducibility, a reasonable narrowing of the DIBH gating window may be implemented with minor changes to the institutional protocol. The observed trend highlights the potential for improved consistency with the experimental approach and supports further investigation to better understand the underlying factors and strengthen these findings in future studies.

## 1. Introduction

Hypofractionated radiotherapy regimens are becoming the standard for adjuvant breast irradiation because of their ability to reduce the overall treatment time without compromising tumor control [1, 2, 3]. For instance, a schedule of 40 Gy delivered in 15 fractions, commonly referred to as the control arm is widely accepted [4, 3]. However, the increased adoption of more compact schedules, such as 26 Gy in 5 fractions commonly referred to as the experimental arm (one-week hypofractionated schedules) [1, 3, 5], has raised concerns regarding cardiac radiation dose [6, 7]. This is particularly concerning for patients with left-sided breast cancer, as they are more susceptible to cardiac toxicity.

An essential approach is to adopt Deep-Inspiration Breath-Hold (DIBH), which is an important respiratory motion-management technique that reduces cardiac toxicity by expanding the thoracic cavity and displacing the heart from the target volume [8, 9]. However, the dynamic nature of the thoracic region during respiration is complex, the breast, heart, and lungs undergo multidimensional and nonlinear movements [9]. Hence, quantifying the radiation dose to the heart is essential. Although a direct approach is difficult to achieve, indirectly, anterior-posterior movement of the lung predicts it best [10].

A key aspect in hypofractionated radiotherapy is ensuring intra-fraction stability and inter-fraction reproducibility to maintain consistent breath-hold positioning for accurate dose delivery and optimal cardiac sparing, both within and across all treatment sessions. Variations in DIBH amplitude within and across fractions can introduce geometric uncertainties, leading to inconsistent target coverage and increased dose to organs at risk. Therefore, determining a patient-specific respiratory amplitude threshold is crucial [11]. Although DIBH is a well-established respiratory motion management technique, rigorous quantification of intra-fraction stability and inter-fraction reproducibility during the beam-on phase remains necessary. While gating thresholds are typically defined by institutional protocols, detailed analysis of the respiratory waveform might allow refinement and potential narrowing of the gating window on an individual basis, which is particularly important in hypofractionated regimens where a reduced number of fractions amplifies the impact of variability. This study stands out for its unique comparison of the stability and reproducibility of the DIBH procedure across two treatment regimens: moderate-hypofraction and a one-week hypofraction Schedule.

Considering these factors, this study aims to systematically investigate the reproducibility and stability of DIBH patterns in the context of hypofractionated radiotherapy. Precisely, we make the following contributions in this work,

- We performed a large-scale quantitative analysis of Varian RPM respiratory waveforms on 510 leftsided breast cancer patients enrolled in the HYPORT Adjuvant trial. The intra-fraction stability and inter-fraction reproducibility of DIBH amplitudes during beam delivery were comprehensively assessed. Throughout this manuscript, stability and intra-fraction stability are used synonymously, as are reproducibility and inter-fraction reproducibility.
- We present a comparative analysis between a moderate hypofractionated (control arm) and a one-week hypofractionated (experimental arm) radiotherapy schedules to investigate the robustness of the DIBH process across different fractionation regimens.
- We further analyze the first five fractions of the moderate-hypofractionation arm to specifically investigate the effect of treatment duration and fraction number on DIBH reproducibility.
- We quantify the impact of structured DIBH training by analyzing changes in respiratory amplitude and expansion of volume of air in the lung between free-breathing and breath-hold CT scans.
- We demonstrate that comparable cardiac sparing and breath-hold stability can be achieved across both treatment regimens despite differences in prescription dose and treatment time.

## 2. Methodology

### 2.1. Study Design

The HYPORT Adjuvant trial compares a one-week hypofractionated adjuvant radiotherapy schedule with the three-week hypofractionated regimen in patients undergoing adjuvant treatment for left-sided breast cancer [3, 5]. Figure. 1 presents the CONSORT diagram, including the total number of patients in both arms, center-wise patient distribution, and the numbers of leftsided, right-sided, and bilateral breast cancer cases.

**Figure 1:**
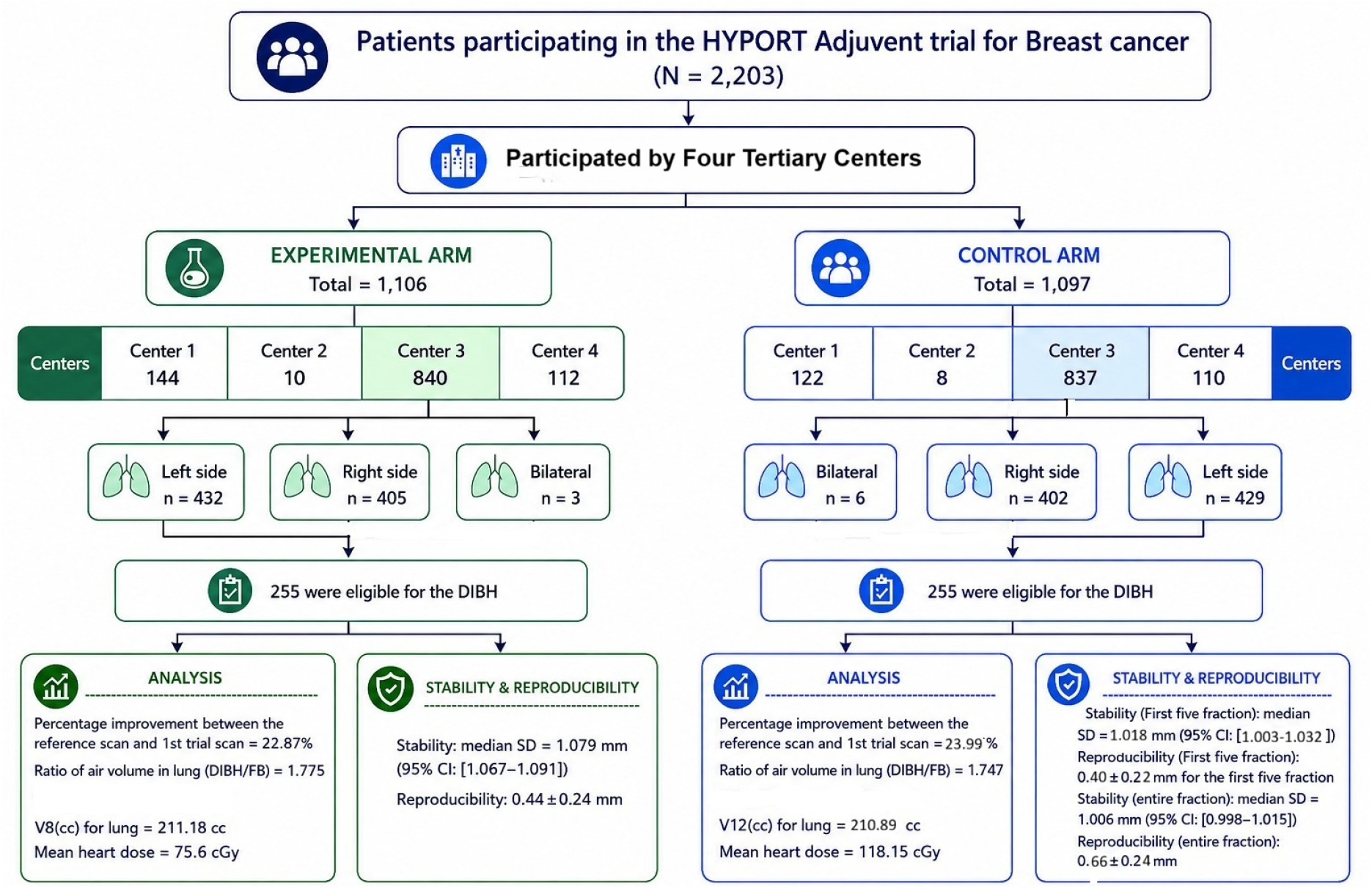
Overview of patient distribution and DIBH outcomes in the HYPORT adjuvant trial. The diagram shows how patients were distributed across centers and by treatment side (left, right, or bilateral). Improvements from the first trial to the reference scan indicate better breath-hold performance and increased air volume in lung, along with corresponding lung and heart dose measures. Stability and reproducibility results demonstrate that patients were able to maintain consistent breath-hold levels throughout treatment in both arms.

Only patients with left-sided breast cancer were considered in this study, as they alone underwent the DIBH process. Patients with bilateral or right-sided breast cancer were excluded since DIBH was not indicated for them. Additionally, left breast cancer patients with internal mammary node (IMN) involvement treated with Tomotherapy^®^, were excluded from the respiratory motion management technique. The remaining left-sided breast cancer patients were treated on a linear accelerator using the DIBH respiratory motion management technique. In each arm, 255 patients met the eligibility criteria for DIBH, resulting in a total cohort of 510 (255 × 2) patients who underwent DIBH training and external beam radiotherapy.

In the control arm, 3812 of 3825 (255 patients × 15 treatment sessions) recorded waveforms (events) were included, and in the experimental arm, 1273 of 1275 (255 patients × 5 treatment sessions) events were retained for analysis. For reproducibility assessment, eight patients in the control arm and three in the experimental arm were excluded because their values exceeded the 5 mm gating threshold. A consistent 5 mm gating window(+3 and -2 mm), defined by the upper and lower thresholds, was maintained for all patients. Occasionally, sudden patient movement or couch shifts resulted in noticeable baseline amplitude drift. Any deviation beyond this predefined window was considered an error, and the corresponding waveforms were excluded from the stability and reproducibility analysis.

Ethical approval for this study was obtained from the Institutional Review Board of Tata Medical Center, Kolkata (approval no. EC/WV/TMC/68/25).

### 2.2. DIBH Training

The DIBH procedure was performed using predefined respiratory gating thresholds to ensure consistent and reproducible breath-hold amplitudes. Respiratory motion was monitored through the vertical displacement of six retroreflective markers positioned on the patient’s Xiphisternum. These markers surrogate diaphragm movement and monitor changes in respiratory amplitude during treatment. Prior to the CT simulation, all patients participated in a standardized DIBH training as part of the HYPORT Adjuvant protocol[3, 5]. The protocol comprised three dedicated training sessions Trial 1, Trial 2, and Trial 3, with the trial 3 session performed during the CT simulation scan. All sessions were typically scheduled 3–4 days apart, to ensure familiarity with the DIBH technique and RPM system. During these sessions, the patients received instructions on achieving and maintaining a stable inspiration breath-hold, as illustrated in (Figure 2).

**Figure 2:**
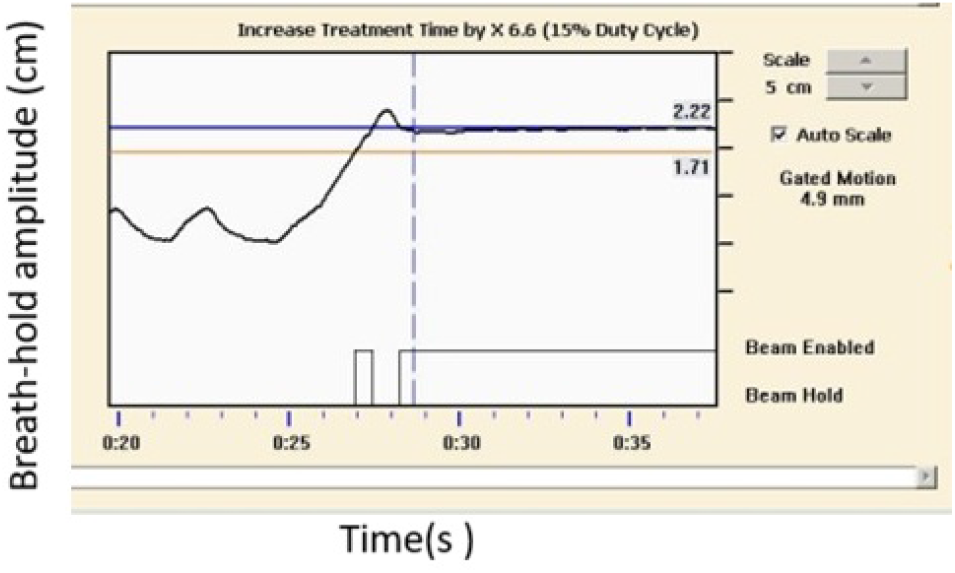
Online breath-hold pattern recorded using the Varian realtime position management (RPM) system. The graph illustrates breathing amplitude over time during an inspiration breath-hold. The upper and lower horizontal lines represent the gating thresholds, and radiation delivery occurs only when the patient maintains a stable breath-hold within the gating window.

For CT simulation during DIBH training, LightSpeed Xtra 16-slice CT simulator (GE Healthcare, Chicago, IL, USA) coupled with the RPM Respiratory Gating System v1.7 (Varian Medical Systems, Palo Alto, CA, USA) was used. The six dot Retro reflective marker placement was individualized for each patient during the first training session and maintained for subsequent trial sessions and stimulation CT scans. Respiratory waveforms recorded during each session were saved as .vxp files.

Planning CT was acquired with a slice thickness of 2.5 mm, including both breath-hold and free-breath scans. An individualized baseline amplitude was established for each patient based on the observed displacement during DIBH. A uniform gating window of 5 mm (− 2 mm to +3 mm relative to the breath-hold amplitude) was applied across all patients to standardize beam delivery conditions.

### 2.3. External Treatment Planning and Delivery

All treatment planning and delivery procedures were performed in accordance with the HYPORT Adjuvant treatment protocol [3, 5]. Treatment planning was performed using the Eclipse treatment planning system (versions 15 and 17, Varian Medical Systems). Intensity-modulated radiotherapy (IMRT) plans were calculated using the Anisotropic Analytical Algorithm (AAA). All treatment plans were created using DIBH CT scans. Dose constraints for both the experimental and control arms were maintained according to the HYPORT adjuvant protocol[3]. Treatment delivery and guidance were performed using a Varian respiratory motion management system.

All treatments were delivered using Novalis Tx linear accelerators (Varian Medical Systems, Palo Alto, CA, USA) and TrueBeam linear accelerators(Varian Medical Systems, Palo Alto, CA, USA). Respiratory waveforms recorded during each treatment fraction were stored as Varian Visitrax playlist (.vxp) files. The DIBH amplitude thresholds during stimulation were maintained throughout the treatment course for each patient.

### 2.4. Respiratory Waveform Analysis Using BreathHold Amplitudes

RPM data were stored in .vxp files, structured into a header and a data section. The header included metadata such as version, data layout, patient ID, date, total study time, sample per second, and scale factor. The data section contains lines corresponding to the data layout fields: amplitude, phase, timestamp, valid flag, TTL(transistor–transistor logic) in, mark, and TTL(transistor–transistor logic) out [12]. In DIBH scans, the amplitude corresponds to the z-coordinate of the respiratory motion.

The RPM camera tracked six reflective markers to establish baseline respiratory patterns. The amplitude data were normalized such that the extreme minimum during free breathing was set to 0 mm, and all subsequent DIBH amplitudes were expressed relative to the baseline. This is illustrated in Figure 3, which represents the waveforms with amplitude (cm) on the Y-axis and timestamps (ms) on the X-axis.

**Figure 3:**
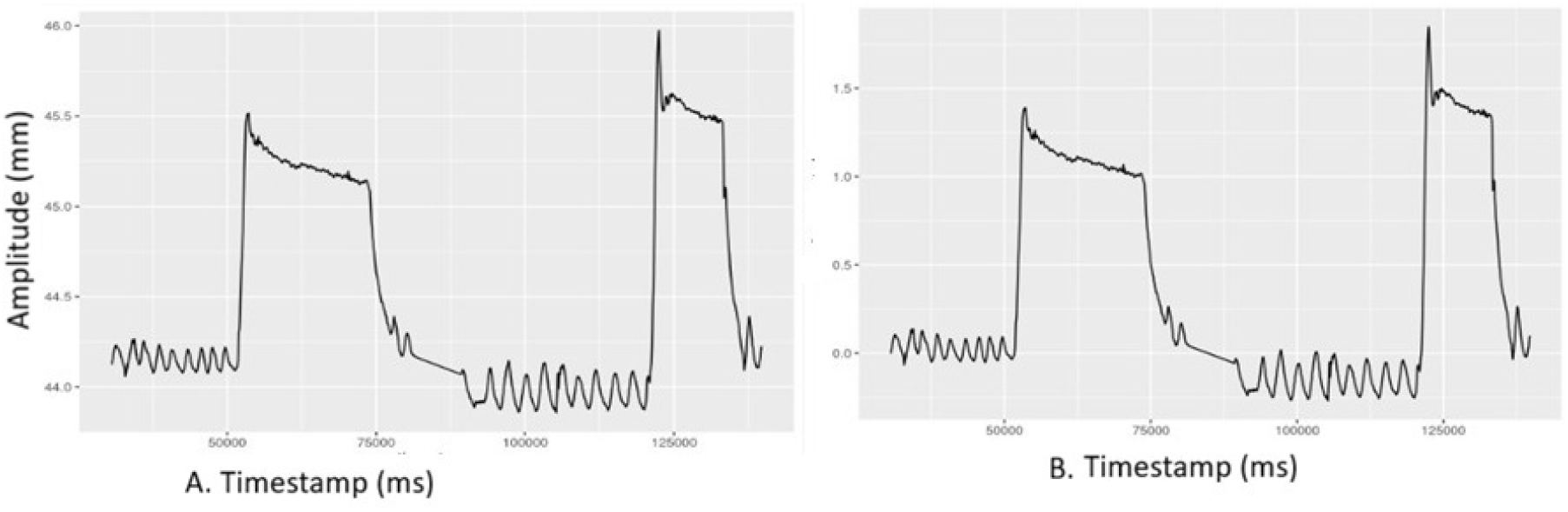
(a) Unnormalized waveform plotted directly from the raw respiratory amplitude.(b) Normalized waveform obtained after setting the extreme minimum value during free breathing to 0 mm and scaling the signal accordingly.

The beam-on time was determined separately for both reference and treatment sessions. Data were extracted using a Python script and converted into longformat variables for subsequent analysis. The processed data were exported as CSV files for further analysis.

For each patient, the upper and lower threshold values were recorded during all DIBH trial sessions. These thresholds defined the breath-hold position and were used to measure the respiratory amplitude achieved during DIBH. The improvement in respiratory amplitude was assessed by comparing the final session with the initial training sessions. The percentage change (PC) in the upper threshold amplitude between successive trial sessions was calculated to quantify the improvement in the consistency of the patient’s breath-hold performance.

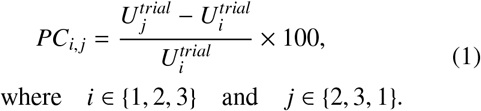

where *i* ∈ {1, 2, 3} and *j* ∈ {2, 3, 1}.

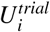 and 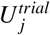 represent the upper threshold of the i^*th*^ and j trial sessions and *PC*_1,2_, *PC*_2,3_, *PC*_3,1_ denote the percentage changes between trial 1, trial 2 and trial 3 respectively. The threshold values from the last training session (simulation scan) were used to set the final gating levels for the treatment sessions. An R script was used to merge the file data and the DIBH training session data, based on patient ID and session number. For each patient, the resulting file was then merged with their corresponding dosimetric data.

### 2.5. Statistical Analysis

We performed a statistical analysis to quantitatively assess the stability and reproducibility of the DIBH waveforms[11]. Here, stability refers to the consistency of the breath-hold amplitude. For each treatment session of a specific patient, it is measured as the Standard Deviation (SD) of the amplitude of breath-hold recorded during the beam-ON period [11]. The 95% confidence intervals of this SD for the two treatment arms were calculated to assess the precision of these estimates. Thereafter, we performed a statistical comparison of the two distributions using appropriate hypothesis testing methods.

For each patient, reproducibility refers to the ability to maintain a consistent breath-hold amplitude during the beam-on periods across each treatment session with respect to the reference amplitude set during the simulation scan. For a patient, we measure it as the average difference between the median breath-hold amplitude in each treatment session and the patient-specific reference (simulation) waveform during the beam-on period. Similarly, for each arm, we compute a histogram of these reproducibility values for all patients (fig. 6).

To determine the clinical relevance of DIBH waveform stability during beam-on, a dosimetric analysis is performed using a uniform treatment planning protocol, to allow consistent comparison of heart and lung doses, as well as treatment delivery time between the two arms. Because lung inflation during DIBH directly affects cardiac displacement and dose distribution, the volume of air in the lung is a key indicator of the quality of breathhold. Therefore, all patients undergo the same DIBH training protocol, and its effectiveness is evaluated by comparing the volume of air in the lung for free-breath and breath-hold CT scans, expressed as mean air volume ratios with 95% confidence intervals (CIs).

## 3. Results

### 3.1. Baseline air volume expansion in lung and DIBH training effect

During DIBH, increased lung inflation elevates intrathoracic air volume, which manifests as a measurable superior displacement of external reflective markers compared with free breathing. This observation was further validated experimentally, demonstrating the mean air volume in the lung during DIBH was substantially higher across all patients in both treatment arms (Table. 1). The mean air volume in lung ratio was 1.775 ± 0.303 for the experimental arm and 1.747 ± 0.285 for the control arm, corresponding to an approximately 1.7-fold increase during DIBH compared with free breathing. The 95% confidence intervals of the experimental arm([1.738, 1.812]) and the control arm([1.712, 1.783]) showed considerable overlap, indicating no meaningful difference in the ratio of expansion of air volume in the lung between the two arms. This is further supported by the minimal difference in mean air volume in the lung ratio ( nearly about 1.7 fold) for both arms, suggesting consistent lung inflation irrespective of the fractionation scheme. Furthermore, the range encompassing 95% of patients confirmed that the increase in lung air volume during DIBH was substantial and consistent throughout the cohort.

**Table 1:** Air volume ratio in Lungs during breath-hold and free breathing for both treatment arms.

| Treatment Arm | Mean | SD | 95% CI | 95% patients |
| --- | --- | --- | --- | --- |
| experimental | 1.775 | 0.303 | [1.738, 1.812] | [1.201, 2.404] |
| control | 1.747 | 0.285 | [1.712, 1.783] | [1.247, 2.336] |

Beyond the expansion of air volume in the lung, the relative vertical displacement of surrogate reflective markers was evaluated as a quantitative indicator of lung expansion during DIBH. This displacement, used to define the optimal gating amplitude, was determined through three consecutive training sessions to ensure reproducibility and patient-specific consistency before treatment delivery. Box plots of all the training session waveforms of all the patients were plotted for both groups of patients, as demonstrated in Figure. 4(a) and (b). A higher proportion of outliers in both arms was observed above the upper quartile rather than below the lower quartile, indicating that several patients achieved breath-hold amplitudes exceeding the population median. This suggests that a subset of patients performed DIBH more effectively than the overall cohort. Moreover, the increased spread observed in the reference session compared to the first trial session reflects inter-patient variability, indicating that improvements in breathing amplitude were not uniform across all patients.

**Figure 4:**
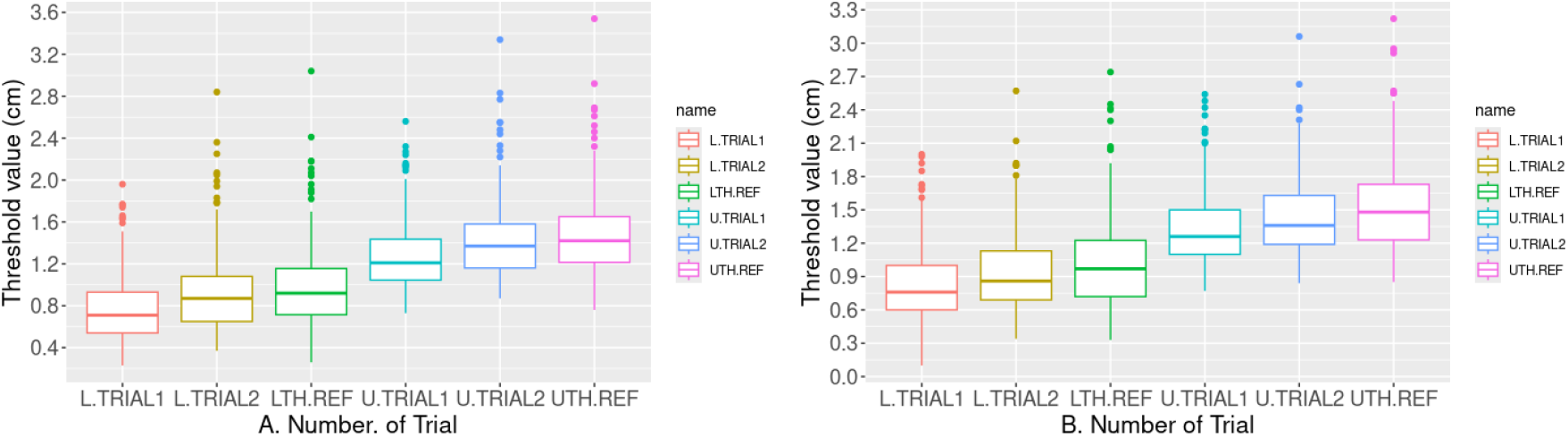
Representative Upper thresholds of (a) Control arm and (b) Experimental arm showing increased spread. Boxplots of the upper threshold recorded during all training sessions for patients in the (a) control arm and (b) experimental arm. Most outliers were observed above the upper quartile, indicating that some patients achieved higher breath-hold amplitudes . The wider spread seen in the reference session compared to the first trial session reflects differences(breath hold amplitude) in training performance among patients.

As illustrated in Table 2, a mean percentage increase in respiratory amplitude was observed from the first trial session to the reference session in both the experimental (22.87%) and control (23.99%) arms, demonstrating the effectiveness of repeated training in improving the DIBH performance. No statistically significant difference was observed between the two arms (p=0.6170), indicating comparable changes in mean percentage increase of respiratory amplitude in both arms. A significant gain in amplitude was observed between the first and second trial sessions in both the experimental (15.02%) arm and control (17.57%) arm.

**Table 2:** Percentage increase of upper threshold amplitude across different trial sessions in experimental and control arms. Statistical comparison between the Experimental arm and the Control arm was performed using p-values, where *p* < 0.05 indicates statistical significance.

| Parameter | mean | SD | 95% CI | P-value |
| --- | --- | --- | --- | --- |
| PC12 (Experimental) | 15.02 | 15.96 | 13.05–17 | 0.1311 |
| PC12 (Control) | 17.57 | 21.53 | 14.91–20.24 |  |
| PC23 (Experimental) | 14.82 | 18.10 | 12.58–17.06 | 0.8241 |
| PC23 (Control) | 14.49 | 15.03 | 12.60–16.39 |  |
| PC31 (Experimental) | 22.87 | 26.36 | 19.61–26.13 | 0.6170 |
| PC31 (Control) | 23.99 | 23.98 | 21.03–26.95 |  |

### 3.2. Dose to Organ at Risk

Low-dose spillage to the lung and heart is a critical determinant of long-term toxicity. Therefore, equivalent dose levels were compared between the treatment arms for the heart and lungs. Low-dose exposure to the equivalent lung volume and the mean heart dose from the treatment planning system were evaluated for both the experimental and the control arms (Table. 3). Specifically, the volume receiving 12Gy equivalent dose to the lung in the control arm was compared with the volume receiving 8 Gy equivalent dose in the experimental arm. These were further analyzed to quantify the benefits of implementing respiratory motion management during treatment planning. The lung volume receiving equivalent doses in the experimental and the control arms was approximately 211 cc in both the arms. The mean heart dose was 75.6 cGy (2.91% of 26 Gy) in the experimental arm and 118.51 cGy (2.95% of 40 Gy) in the control arm, respectively.

**Table 3:** Dosimetric parameters of mean Heart dose and Lung volume summary. For control arm V12(CC) and experimental arm V8 (CC).

| Parameter | Median | SD | 95% CI |
| --- | --- | --- | --- |
| **Experimental arm** |  |  |  |
| Ipsilateral Lung V8 (CC) | 211.18 | 61.58 | 203.58–218.77 |
| Heart mean dose(cGy) | 75.6 | 23.39 | 72.18–77.95 |
| **Control arm** |  |  |  |
| Ipsilateral Lung V12 (CC) | 210.89 | 60.51 | 203.41–218.37 |
| Heart mean dose(cGy) | 118.51 | 40.1 | 113.56–123.45 |

### 3.3. Intra-fraction Stability of Breath-Hold Amplitudes

The stability values over all patients from each treatment arm were considered and the maximum observed values were 1.94 mm and 1.91 mm for the experimental and control arms, respectively. The median value of breath-hold stability for intra-fraction amplitude was 1.006 mm (95% CI: [0.998–1.015]) for the control arm and 1.079 mm (95% CI: [1.067–1.091]) for the experimental arm. For the first five fractions of the control arm, the median value was 1.018 mm (95% CI: [1.003– 1.032]). The corresponding histograms for the control and experimental arms are presented in Figure 5(a) and (b), respectively. Histograms show the number of events plotted against the standard deviation in all treatment sessions for all patients.

**Figure 5:**
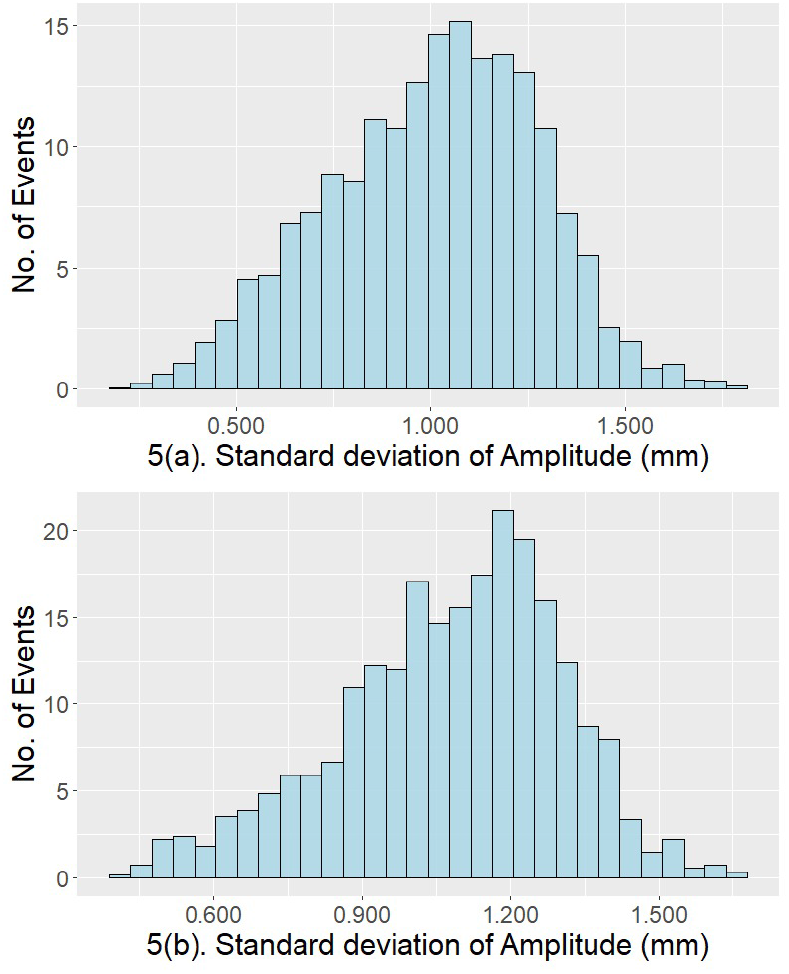
Histograms of the standard deviation of respiratory waveform amplitudes across all treatment events for all patients: (a) Control arm and (b) Experimental arm. The distributions illustrate the variability in breathing amplitude during all treatment sessions for both arms

The Shapiro–Wilk test indicated that the amplitude measurements were not normally distributed in both the experimental arm (W = 0.8896, p < 0.001) and the control arm (W = 0.5765, p < 0.001). Accordingly, nonparametric statistical methods were employed to compare the two arms. The Mann–Whitney U test yielded a p-value of 0.05636, suggesting a trend toward a difference in breath-hold stability between the two arms. However, this difference did not reach statistical significance at the 5% significance level, indicating that the stability of breath-hold was comparable between the experimental and control arms.

### 3.4. Inter-fraction Reproducibility of Breath-Hold Amplitudes

In the experimental arm, the mean deviation from the reference amplitude was (0.44 ± 0.24) mm (95% CI: 0.41–0.47 mm) for 224 patients. A total of 245 out of 252 patients were within a 2 mm deviation from the reference amplitude (Table 4, Figure 6(a)), demonstrating high reproducibility in the experimental arm.

**Table 4:** Distribution across patients of the perpatient average difference 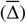 between median DIBH amplitude values from each treatment session and those from the corresponding reference scan for the experimental arm.

| $\Delta$<br>(mm) | No. of<br>patients | Mean<br>(mm) | SD<br>(mm) | 95% CI (mm) | |
| --- | --- | --- | --- | --- | --- |
|  |  |  |  | Lower | Upper |
| 0.0713–1.07 | 224 | 0.44 | 0.244 | 0.41 | 0.47 |
| 1.07–2.07 | 21 | 1.58 | 0.28 | 1.45 | 1.70 |
| 2.07–3.07 | 5 | 2.45 | 0.34 | 2.15 | 2.74 |
| 3.07–4.07 | 2 | 3.65 | 0.26 | 3.29 | 4.01 |

**Figure 6:**
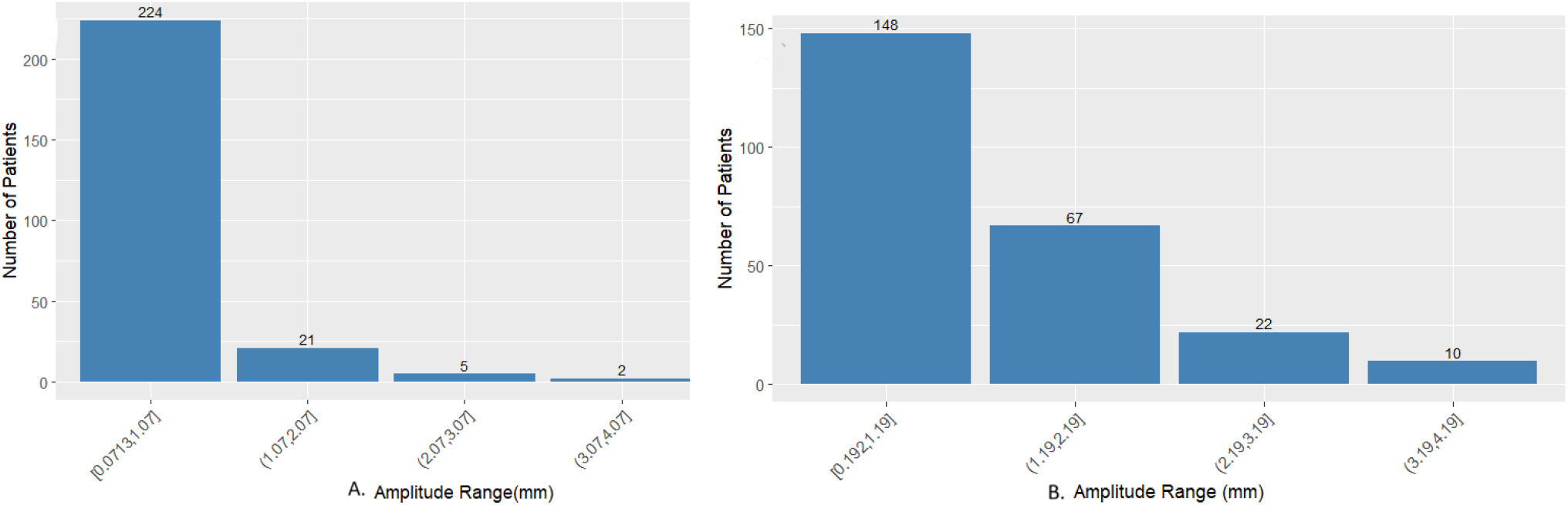
Histogram plot of patient amplitude range with the number of patients in (a)Experimental arm.(b)control arm.These plots represent the interfraction reproducibility of breath-hold amplitudes relative to the simulation scan across patients in each treatment arms.

For the control arm, the mean deviation from the reference scan was (0.66 ± 0.24) mm (95% CI: 0.62–0.70 mm), as presented in Table. 5 and Figure. 6(b). A total of 58% (n = 148/255) of the fractions exhibited stable amplitude variations within the range of 0.19–1.19 mm. The amplitude exhibited greater variation, extending up to 2.19 mm, with 84.3% of patients within this range. On the contrary, the experimental arm showed a smaller average deviation and a higher percentage of reproducibility in breath-holds compared to that of the control arm.

To examine whether the improved outcomes in the experimental arm could be due to chance, the reproducibility of the first five fractions of the control arm was analyzed separately and compared with that of the experimental arm. This comparison assessed the inter-fraction reproducibility during the initial treatment phase (i.e., the first 5-fractions) of the control arm relative to the complete treatment course of the experimental arm. As shown in Table 6, the average deviation between the reference breath-hold amplitude and the first five fractions of the control arm was 0.40 ± 0.22 mm (95% CI: 0.37–0.43 mm), which is close to the value observed in the experimental arm (0.44 ± 0.24 mm, 95% CI: 0.41–0.47 mm). This suggests that during the initial treatment phase, the control arm can achieve reproducibility comparable to that of the experimental arm. However, over the full treatment course, the average deviation in the control arm increases to 0.66 ± 0.25 mm (Table 5), indicating that reproducibility may decline with increasing treatment fractions.

**Table 5:** Distribution across patients of the per-patient average difference between median DIBH amplitude values from each treatment session and those from the corresponding reference scan for control arm.

| $\Delta$<br>(mm) | No. of<br>patients | Mean<br>(mm) | SD<br>(mm) | 95% CI (mm) | |
| --- | --- | --- | --- | --- | --- |
|  |  |  |  | Lower | Upper |
| 0.192–1.19 | 148 | 0.66 | 0.24 | 0.62 | 0.70 |
| 1.19–2.19 | 67 | 1.55 | 0.26 | 1.49 | 1.62 |
| 2.19–3.19 | 22 | 2.57 | 0.31 | 2.43 | 2.70 |
| 3.19–4.19 | 10 | 3.51 | 0.25 | 3.35 | 3.67 |

**Table 6:** Distribution across patients of the per-patient average difference between median DIBH amplitude values from each treatment session and those from the corresponding reference scan for first five fraction for control arm.

| $\Delta$<br>(mm) | No. of<br>patients | Mean<br>(mm) | SD<br>(mm) | 95% CI (mm) | |
| --- | --- | --- | --- | --- | --- |
|  |  |  |  | Lower | Upper |
| 0.056–1.06 | 207 | 0.40 | 0.22 | 0.37 | 0.43 |
| 1.06–2.06 | 30 | 1.38 | 0.22 | 1.31 | 1.47 |
| 2.06–3.06 | 11 | 2.31 | 0.25 | 2.16 | 2.46 |
| 3.06–4.06 | 4 | 3.53 | 0.32 | 3.22 | 3.85 |

### 3.5. Treatment delivery time

Both arms demonstrated an average simulation scan time of 13 seconds. In the control arm, the median total treatment time per session was 196 seconds, while treatments that involved tumor bed boost were 262 seconds. In contrast, the experimental arm showed a median treatment time of 381 seconds per session, which increased to 518 seconds when tumor bed boost was included. During the treatment delivery, the dose rate was kept constant for all patients.

## 4. Discussion

This study quantitatively analyzed the variability in DIBH breathing patterns in patients with left-sided breast cancer who underwent radiotherapy in the HYPORT Adjuvant trial at one of the tertiary centers[5]. Both arms showed an approximately 1.7-fold increase in breath-hold air volume in the lung compared to free breath with equal, adequate, and structured training methodology adapted for both arms. The progressive improvements in amplitude observed in the breath-hold technique during the training sessions demonstrated an initial rapid gain of approximately 15–17% between the upper threshold of the first and the second trial sessions. The change in amplitude between the second and the final trial sessions showed only a modest increase. Accordingly, patients were simulated during the third or the final trial session. These findings are in agreement with the study by Jiang et al. [13], who reported that 95.8% of patients were able to maintain stable breathhold by the final day of coaching[13]. In the present study, standard treatment planning techniques and dose constraints were achieved successfully for the HYPORT Adjuvant Trial, with equivalent low lung dose exposure and mean heart doses for both arms, consistent with the prescribed doses. Overall, these results highlight that structured patient education and repeated practice improved DIBH stability and reproducibility, while maintaining comparable intra-fraction stability during beamon periods in both treatment arms.

The stability of breath-hold amplitudes remained within 2 mm for both arms (1.1 mm for experimental and 1.03 mm for control arm), consistent with findings reported by Reitz et al. [11], who observed a median stability of 0.3 mm and a maximum deviation below 2 mm in breath-hold position during beam delivery in DIBH procedure. For inter-fraction reproducibility, the experimental arm showed a statistically significant reduction in average deviation from the reference breathhold amplitude (0.44 ± 0.24mm) compared to the control arm (0.66 ± 0.25 mm) [11]. Although stability was comparable between the two arms, reproducibility differed, with variations in amplitude observed in both. A root cause analysis of this difference was conducted to determine whether it is a systematic or random effect. Therefore, a comparative analysis between the stability and reproducibility of DIBH amplitude during the first five treatment fractions of the control and experimental arms was performed. The intra-fraction stability over the first five fractions in the control arm demonstrated a median value of 1.04 mm, while reproducibility was 0.4 mm, closely aligning with the corresponding results observed in the experimental arm(stability:1.006 mm and reproducibility: 0.44 mm ). These results imply that the apparent discrepancies in reproducibility across the full treatment course are unlikely to originate from the early fractions and may instead emerge over subsequent treatment sessions.

Furthermore, the observed similarity in intra-fraction stability was evaluated in relation to beam-on time during treatment delivery. Although the experimental arm involved longer treatment durations due to the 1 week hypo fractionation dose regimen, amplitude stability during the beam-on period showed no observable dependency on the increased treatment time, reinforcing the robustness of the stability of DIBH, irrespective of the delivery duration.

The consistency in breath-hold stability and reproducibility across treatment sessions suggests that reducing the gating window for DIBH patients from 5 mm (i.e., {+3, −2} ) to 4 mm (i.e., {+2, −2} ) is a realistic step forward. Session-to-session variation was minimal, and respiratory motion remained predictable enough that a narrower window is unlikely to trigger additional gating errors or disrupt treatment workflow, while maintaining the same dose to OAR.

## 5. Conclusion

This study estimated the intra-fraction stability and inter-fraction reproducibility of DIBH in left-sided breast cancer patients undergoing moderate (control arm) and one-week (experimental arm) hypofractionated radiotherapy regimens within the HYPORT Adjuvant trial. In this study, we thoroughly analyzed the respiratory amplitude for both arms during treatment delivery to evaluate the efficacy of the respective DIBH treatment schedules. The present findings confirm the effectiveness of structured DIBH training for both study arms within the HYPORT adjuvant Trial.

The experimental arm of the HYPORT adjuvant trial demonstrated excellent breath-hold stability and reproducibility while consistently meeting the prescribed dosimetric constraints for organs at risk. Both arms exhibited minimal variation in breath-hold amplitude during DIBH, indicating robust intra-fraction stability, despite the longer beam-on time in the experimental arm. The intra-fraction stability of breath-hold amplitude was maintained throughout the treatment course and was not affected by the differing treatment durations of the 5fraction and 15-fraction schedules. Furthermore, analysis of the first five fractions in the control arm revealed stability and reproducibility comparable to those in the experimental arm, while demonstrating superior interfraction reproducibility throughout the treatment schedule.

Collectively, these results provide strong evidence for the clinical reliability and efficiency of the experimental DIBH schedule in the HYPORT Adjuvant study. The improved reproducibility observed throughout treatment supports its suitability for patients with left-sided breast cancer and its broader clinical adoption. Furthermore, the observed stability and reproducibility suggest that the gating threshold window may be safely reduced to ±2mm, although further quantitative validation may be required to confirm this adjustment.

## Data Availability

The data that support the findings of this study are
available from the corresponding author upon reason-
able request. Due to privacy considerations and ethical
requirements related to participant confidentiality, the
data are not publicly available.

## 6. Acknowledgments

## Ethical Approval

The study has been approved by the Institutional Review Board of Tata Medical Center (Approval No. EC/WV/TMC/68/25).

## Data Access Statement

The data that support the findings of this study are available from the corresponding author upon reasonable request. Due to privacy considerations and ethical requirements related to participant confidentiality, the data are not publicly available.

## Author Contributions

**Anurupa Mahata** is responsible for designing and implementing the research, originated the main concept and oversaw the project.

